# Analytical Sensitivity of the BOSCH Vivalytic Molecular Assay for Detection of Monkeypox Virus Clade Ib

**DOI:** 10.64898/2026.01.07.25342807

**Authors:** Camille Escadafal, Kenneth Adea, Placide Mbala, Isabella Eckerle

## Abstract

We evaluated the BOSCH Vivalytic MPXV assay using serial dilutions of monkeypox virus clade Ib, a new offshoot of clade I identified in 2023. The assay detected viral DNA down to ∼100 copies/mL demonstrating comparable analytical sensitivity to our in-house reference PCR and to other commercial platform-based mpox molecular assays.

## Background

Monkeypox virus (MPXV), a double-stranded DNA virus in the genus *Orthopoxvirus*, causes mpox disease in humans. Following the 2022 multi-country outbreak of clade IIb MPXV, a novel variant, clade Ib, emerged in late 2023 in the Democratic Republic of the Congo (DRC). This variant has been responsible for sustained human-to-human transmission across multiple provinces and neighboring countries. MPXV clade Ib demonstrates specific genetic features compared to other MPXV clades including deletions in several genes (1,2). While these can lead to altered transmission and pathogenicity, they may also impact the performance of diagnostic tools. Rapid and accurate diagnostics are essential for outbreak control, yet few molecular assays have been evaluated against clade Ib. While conventional real-time PCR remains the gold standard, its use in decentralized or resource-limited settings is constrained by infrastructure and personnel requirements. Automated cartridge-based molecular platforms can bridge this gap by enabling reliable near point-of-care (near-POC) detection.

## The study

The BOSCH Vivalytic system is an integrated molecular platform for nucleic acid extraction, amplification, and detection in a self-contained cartridge which requires minimal handling from sample to result. The BOSCH MPXV test is fully automated qualitative real-time PCR for detection of monkeypox virus (MPXV) and orthopoxvirus (OPXV) species in COPAN eNAT® transport medium.

We evaluated the Vivalytic MPXV assay’s analytical sensitivity using a cultured isolate of MPXV clade Ib provided by WHO Biohub (catalogue reference 2024-WHO-LS-003) and originating from DRC. Viral propagation was performed using Vero-E6 cells (ATCC CRL-1586), which were cultured in DMEM GlutaMAX I medium supplemented with 10% FBS, 1x non-essential amino acids, and 1% penicillin-streptomycin (all reagents from Gibco, USA). The supernatant was harvested for viral stock preparation. All experiments involving live, infectious MPVX were approved and adhered to the standard operating procedures of our Biosafety Level 3 (BSL-3) facility. The resulting cultured virus was sequenced and uploaded in Genbank under accession number PX049155. Viral DNA quantification was performed using quantified DNA standards and an in-house qPCR targeting the A30L open reading frame (3,4). Serial 1:3 dilutions starting at 3.07×10^4^ DNA copies/mL were prepared in phosphate-buffered saline (PBS). The BOSCH Vivalytic MPXV assay was performed according to the manufacturer’s instructions except for the choice of sample matrix. Instead of a clinical swab in eNAT transport medium, we mixed each viral dilution with an equal volume of eNAT medium. PBS mixed 1:2 with eNAT medium served as a negative control. Sample loading was performed as per the manufacturer’s instructions, adding 300µl of sample into the cartridge. Each test was performed in duplicate.

## Results and Implications

The Vivalytic MPXV assay detected MPXV clade Ib down to 88 DNA copies/mL across both targets (MPXV/OPXV) and both duplicates (Table 1). At a concentration of 40 copies/mL, only one of the duplicates tested positive and the Ct value was over 40. These data support an estimated analytical limit of detection (LoD) of approximately 100 copies/mL for clade Ib under our study conditions.

**Table 1.** Analytical testing results with clade Ib viral dilutions for the BOSCH Vivalytic MPXV assay.

| Dilution | DNA concentration in sample tested (copies/mL) | Cq value – MPX target |  | Cq value – OPX target |  |
| --- | --- | --- | --- | --- | --- |
|  |  | Duplicate 1 | Duplicate 2 | Duplicate 1 | Duplicate 2 |
| 1 | 3.07E+04 | 32.7 | 32.8 | 32.6 | 32.7 |
| 2 | 8.35E+03 | 34.7 | 36.3 | 34.5 | 35.8 |
| 3 | 3.84E+03 | 35.6 | 35.3 | 36.5 | 35.8 |
| 4 | 9.28E+02 | 37.5 | 38.2 | 38.2 | 39.0 |
| 5 | 8.85E+01 | 39.0 | 39.7 | 39.3 | 39.5 |
| 6 | 4.04E+01 | neg | 45.0 | 42.5 | neg |
| 7 | 3.17E+01 | 43.1 | neg | neg | neg |
| 8 | 1.06E+01 | neg | neg | neg | neg |
| Neg | 0 | neg | neg | neg | neg |

This analytical sensitivity is comparable to, or better, than that reported for other manual or near-POC molecular assays for MPXV (5,6)(ref preprint). The LoD is also well below typical viral loads in lesion swabs from confirmed mpox cases (>10^6^ copies/mL) (7). Therefore, the assay is likely to reliably detect clinically relevant, infectious samples.

The Vivalytic platform’s minimal handling, integrated workflow, and rapid turnaround time of less than 60 minutes, make it a potentially suitable tool for decentralized testing in outbreak settings. However, it is a low-throughput system, as each device can perform only one test at a time. Moreover, as it is not battery-powered and requires stable power and controlled environment, it is more appropriate for regional laboratories than remote field use.

One limitation of this study is that it included only MPXV clade Ib and not virus isolates of the other clades (Ia, IIa and IIb). Additionally, this evaluation focuses solely on the analytical sensitivity of the test but not specificity, and may not reflect its clinical performance. In fact, working with cultured virus in a controlled laboratory setting differs from testing clinical specimens in real-life conditions. Future studies should investigate the clinical sensitivity and specificity on lesion swabs from different regions and MPXV clades, in order to confirm the potential for wider use of this assay. In outbreak-prone regions with limited access to laboratories and trained staff, the deployment of cartridge-based molecular platforms could reduce diagnostic delays, improve patient management and strengthen surveillance.

In conclusion, these findings represent the first laboratory evaluation of the BOSCH Vivalytic MPXV assay against MPXV clade Ib. The assay detected MPXV clade Ib with an analytical sensitivity of less than 100 DNA copies/mL. These findings support its potential to enable decentralized molecular testing for mpox in resource-limited settings, thereby improving our ability to detect and respond to outbreaks.

## Data Availability

All data produced in the present work are contained in the manuscript.

## Acknowledgments

We thank BOSCH for lending the Vivalytic platform and providing the MPXV assay cartridges. This research was conducted with biological material - MPXV clade Ib isolate, obtained through the WHO BioHub System, from the WHO BioHub Facility: Spiez Laboratory Switzerland. We wish to gratefully acknowledge the DRC’s Institut National de Recherche Biomedicale (INRB), who originally provided the biological material to the WHO BioHub System. We gratefully acknowledge Francisco PEREZ RODRIGUEZ, Adriana RENZONI, Valentin CHUDZINSKI and Florian LAUBSCHER of the University Hospitals of Geneva, for their contribution to the reference PCR protocol and the sequencing and analysis of the MPXV clade Ib cultured virus.

